# Genomic Analysis of Trichotillomania

**DOI:** 10.1101/2025.01.23.25321045

**Authors:** Matthew W. Halvorsen, Melanie E. Garrett, Michael L. Cuccaro, Allison E. Ashley-Koch, James J. Crowley

## Abstract

Trichotillomania (TTM) is a psychiatric condition in which people feel an overwhelming urge to pull out their hair, resulting in noticeable hair loss and significant distress. Twin, family and candidate gene studies suggest that TTM is at least partly genetic, but no genome-wide analyses have been completed. To fill the gap in this field, we have conducted a case-control study of genotype array data from 101 European ancestry TTM cases and 488 ancestry-matched unaffected controls. TTM cases were ascertained in the USA through web-based recruitment, patient support groups, and conferences organized by the Trichotillomania Learning Center. Following clinical confirmation of a TTM diagnosis, patients completed self-report assessments of frequency and duration of hair pulling, other psychiatric symptoms, and family history. Unaffected controls were also ascertained in the USA and were matched to cases by ancestry. In the first formal genome-wide association study of TTM, we did not identify any common variants with a genome-wide significant (P < 5×10^−8^) association level with case status. We found that TTM cases carry a higher load of common polygenic risk for psychiatric disorders than unaffected controls (*P* = 0.008). We also detected copy number variants previously associated with neuropsychiatric disorders in TTM cases (specifically, deletions in *NRXN1, CSMD1*, and 15q11.2). These results further support genetics’ role in the etiology of TTM and suggest that larger studies are likely to identify risk variation and, ultimately, specific risk genes associated with the condition.

## Introduction

Trichotillomania (TTM) is a psychiatric condition characterized by recurrent pulling out of one’s own hair, leading to hair loss and marked functional impairment.^1^ It affects ~1-2% of the population and is often comorbid with obsessive-compulsive disorder (OCD), anxiety/depressive disorders and attention-deficit hyperactivity disorder (ADHD).^2^ TTM typically begins in childhood and affects males and females equally, though fewer males seek treatment.^2^ DSM-V classifies it in the Obsessive-Compulsive and Related Disorders chapter, and treatment generally involves psychoeducation, habit reversal therapy, and/or medication (e.g., N-acetylcysteine or olanzapine).^1^

The evidence suggests that TTM has a genetic component. Several case reports^3–5^, five family studies^6–10^, and one twin study^11^ all support a role for genetics in the etiology of TTM. Candidate gene studies in humans have described rare coding variants in *SLITRK1*^*12*^ and *SAPAP3*^*13*^ associated with TTM. Furthermore, in mice, single-gene knockouts of multiple genes have been shown to induce compulsive grooming (e.g., *Sapap3*,^14^ *Slitrk5*^*15*^ and *Hoxb8*^*16*^). The literature lacks an agnostic genome-wide analysis of TTM in humans using modern genomic methods. Here, we apply such methods to 101 TTM cases and 488 matched controls. See ***Figure 1*** for an overview of this study.

**Figure 1.**
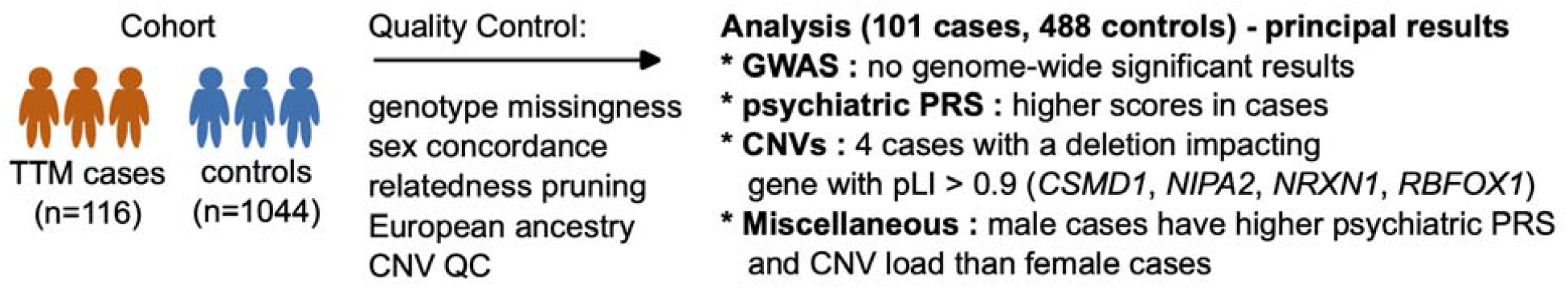
Overview of the case/control study. We collected genotype array data from 116 TTM cases and 1044 unaffected controls and, after rigorous quality control, identified a subset of 101 cases and 488 controls suitable for comparison. We provide an overview of the principal results for this study, which are consistent with 1) genetic contribution to TTM risk and 2) contributions to risk from both common and rare genetic variation.

## Material and methods

### Ethics

This study was carried out in accordance with the latest version of the Declaration of Helsinki and was approved by the Institutional Review Board (IRB) of Duke University Medical Center. All individuals provided written informed consent.

### Cases

Families with at least one TTM case were ascertained primarily through web-based recruitment and patient support groups, both in the United States. Our clinical team also recruited subjects by attending Trichotillomania Learning Center (TLC) conferences. Using all available clinical data, including prior diagnosis, affection status was derived using DSM-IV criteria. All cases were adjudicated by a board-certified psychiatrist and licensed clinical psychologist with extensive experience in the diagnosis and treatment of TTM. Once enrolled, self-reports of frequency and duration of hair pulling, other psychiatric symptoms and family history were obtained. Self-reports of anxiety, depression, and OCD were obtained on adult subjects through the State-Trait Anxiety Inventory (STAI-Y), Beck Depression Inventory Second Edition (BDI-II), and Obsessive-Compulsive Inventory (OCI) questionnaires, respectively. Family history was collected through the proband unless the proband was a minor. See the ***Supplement*** for details and results from these self-report measures.

### Controls

Control subjects from the United States were collected by a marketing research company, Knowledge Networks (Menlo Park, Calif.), as part of the Molecular Genetics of Schizophrenia Part 2 study.^17^ Knowledge Networks recruited controls by random-digit dialing of residential phone numbers. Participants were pointed to a website to learn more about the study, given preliminary online informed consent, and completed a self-report clinical assessment. The online assessment included the Composite International Diagnostic Interview-Short Form (CIDI-SF), modified for lifetime common mood, anxiety, and psychosis or bipolar disorder. We excluded individuals who failed to complete the CIDI-SF or endorsed schizophrenia, schizoaffective disorder, auditory hallucinations, delusions, or bipolar disorder.

### Genetic data

For cases, DNA from 116 unrelated TTM cases were genotyped on an Illumina GSAv2 array, which consists of 665,608 SNP markers in total. Genotyping occurred at the SNP&SEQ Technology Platform at Uppsala University, Sweden. For controls, genotype array data from 1044 matched controls was kindly shared by Dr. Patrick Sullivan at UNC Chapel Hill. These samples were genotyped on an Illumina array that featured a backbone of GSAv2 content, with additional markers added (805,379 markers total).

### Genomic analyses

Detailed methods for all genomic analyses are presented in the ***Supplement***. In brief, we first merged case and control genotype data for overlapping markers and performed extensive pre-analysis quality control (QC). This was followed by genotype imputation and a genome-wide association study (GWAS) of TTM. Next, for each case and control individual, we calculated polygenic risk scores (PRS) for a composite measurement of psychiatric disorder polygenic risk, derived from a cross-disorder study of 8 major psychiatric disorders from the Psychiatric Genomics Consortium (PGC) Cross-Disorder Working Group.^18^ We also calculated PRS for one negative control phenotype (standing height^19^). We hypothesized that TTM cases would have elevated PRS for psychiatric disorders but not standing height. This was followed up by calculating PRS for three additional phenotypes: OCD,^20^ ADHD,^21^ and major depressive disorder (MDD).^22^

We also used the genotype array data to call copy number variants (CNVs; described in the ***Supplement***), which are large deletions or duplications of DNA sequences. We compared the burden of large, rare, deleterious CNVs, hypothesizing that cases would carry a higher burden of these variants impacting genes defined as ‘constrained’ in the general population.

## Results

### TTM sample characteristics

Our TTM cases are primarily female (91%), Caucasian (89%), and were young at the time of enrollment (mean age of 28.8 years, with a range of 7-60 years). The mean age at onset for all probands was 12.1 years (12.4 years in females and 9.6 years in males). As detailed in ***Tables S1-S5***, many cases reported comorbid psychiatric conditions (e.g., anxiety, OCD, depression). The frequency of self-reported TTM in first-degree relatives of cases (10.7%) exceeded contemporary estimates of TTM population prevalence (~1-2%).^2^ This was also true for a composite set of body-focused disorders (nail biting, skin picking, hair pulling/twirling, lip biting, body dysmorphic disorder (BDD), and obsessive/compulsive grooming), which were endorsed by 33.7% of first-degree relatives (***Table S1***).

### Genome-wide association study

We compiled a genome-wide association study (GWAS) of common variation within a European-ancestry cohort of 101 TTM cases and 488 controls (***Figure S1***). The summary statistics did not display any evidence of test statistic inflation (lambda = 1.011, ***Figure S2***). As might be anticipated, given the sample size and what has been seen in GWAS of other psychiatric cohorts, there were no loci associated with TTM at a genome-wide level of significance (P < 5×10^−8^, ***Figure S3***).

### Case/control comparison of polygenic risk scores

Given that the GWAS did not provide evidence for TTM loci, polygenic risk scores (PRS) were used to assess whether there was a common genetic liability for TTM. Our dataset is underpowered for use in SNP-based heritability estimation, and, to our knowledge, there is no other TTM GWAS with which we can compare our summary statistics. Instead, we tested the hypothesis that some amount of TTM genetic risk is pleiotropic with shared genetic risk for other psychiatric disorders. For this, we computed a cross-disorder PRS (using GWAS summary statistics from the Psychiatric Genomics Consortium (PGC) Cross-Disorder Working Group^18^) for our samples and compared the mean PRS in cases to controls. We utilized a PRS for standing height as a negative control, since this trait is not thought to run orthogonal to psychiatric risk.

TTM cases showed elevated PRS for risk of a psychiatric outcome relative to controls (estimate = 0.310, P=0.008, ***Figure 2***), but no difference in PRS for standing height (estimate = −0.028, P=0.808). We also tested for a difference in PRS for specific psychiatric disorders (OCD,^20^ ADHD^21^ and MDD^23^) between TTM cases and controls. These disorders were selected based on high comorbidity with TTM. No evidence was found for a case/control difference in PRS (P<0.05) for any of these three disorders (not shown). After applying a Bonferroni correction for these five tests, the adjusted p-value for cross-disorder psychiatric outcome is still significant (adjusted P=0.04). We used the first genetic principle component (PC1) and sex as covariates in all five tests since both were associated with case-control status. To ensure that samples with extreme PC1 values weren’t driving this difference, we repeated standing height and psychiatric case PRS comparisons using only samples with PC1 < 0.05 (case/control N = 80/457). The initial observations held in this subset, with no difference in PRS for standing height (estimate=-0.114, P=0.359), and a significant difference in psychiatric case PRS (estimate=0.382, P=0.003).

**Figure 2.**
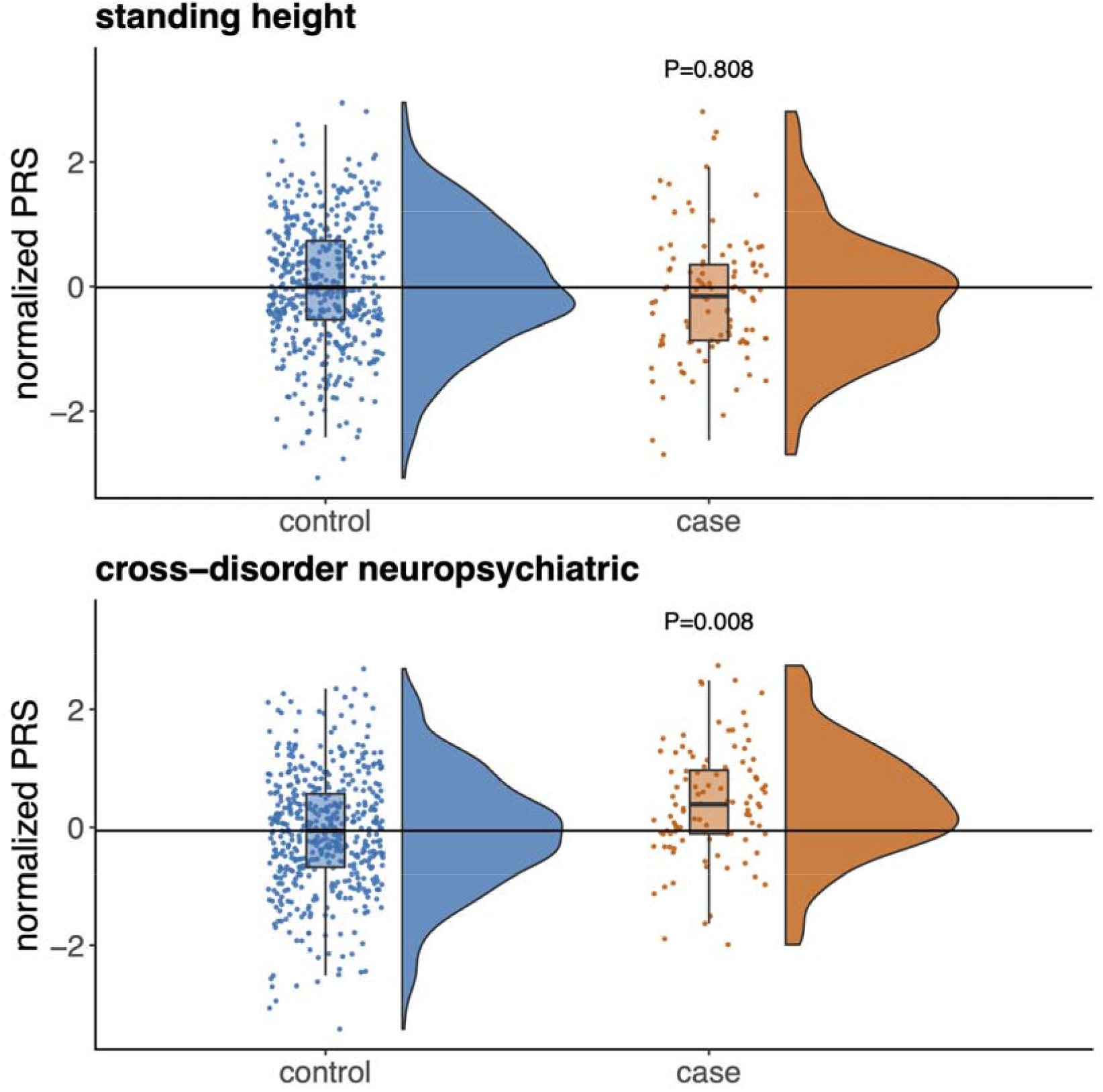
Comparisons of polygenic risk scores (PRS) in 101 European TTM cases versus 488 population-matched controls. Results from comparison of normalized PRS for standing height^19^ and neuropsychiatric case status.^18^ There is no detectable difference in height PRS between cases and controls (top). Still, there is a significant case/control difference for psychiatric PRS (0.310, P=0.008).

We noted a trend for a difference in psychiatric PRS risk between male TTM cases (n=12) versus female (n=89). In comparisons with sex-matched controls, both displayed the same levels of evidence for a difference versus controls (two-sided T-test P=0.031 and 0.012 for male and female comparisons, respectively). However, the point estimate of the difference in PRS versus controls appeared noticeably higher in males (normalized PRS difference of 0.81 vs 0.21, ***Figure S4***).

### Case/control comparison of CNV burden

We next compared the burden of large (>100kb, >=15 markers), deleterious CNV deletions and duplications in TTM cases and controls, testing the hypothesis that cases carry a higher burden of these variants than controls, specifically within genes defined as ‘constrained’ in the general population (probability of loss-of-function intolerance (pLI) > 0.9 in gnomAD v2.1.1).^24,25^ Statistical power was limited due to our sample size, but we did note a trend toward an increased burden of deletions in constrained genes in cases (OR=3.33, P=0.09). Of the case deletions impacting constrained genes (four total, ***Figure 3***), one impacts a known neurodevelopmental gene (*NRXN1*), one overlaps the coding region of a novel neurodevelopmental gene (*CSMD1*^*26*^), one is a known neurodevelopmental CNV (15q11.2), and another deletes a substantial fraction of *RBFOX1*, which is primarily expressed in brain. Plots of intensity data underlying all four calls are provided in ***Figure S5*** and ***Figure S6***.

**Figure 3.**
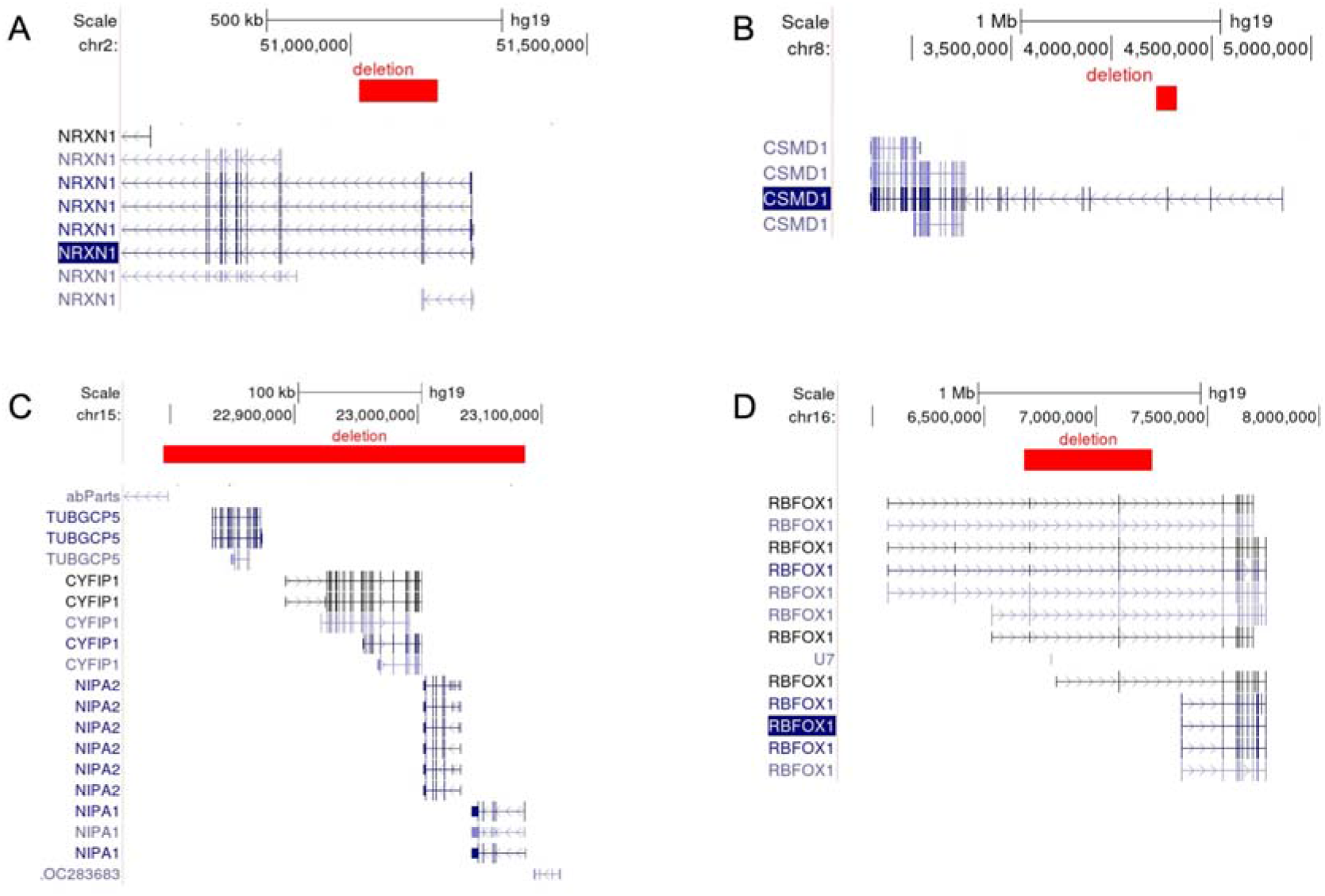
Deletions in TTM cases overlapping a gene with pLI>0.9. All genomic coordinates provided for breakpoints correspond to human reference build 37. A) Deletion disrupting *NRXN1* (chr2: 51017206 - 51183559). B) Deletion that removes part of *CSMD1* (chr8: 4224080 - 4327992). C) A 15q11.2 deletion (chr15: 22794494 - 23086929). D) Deletion overlapping a substantial portion of *RBFOX1* (chr16: 6680132 - 7253942).

We also note a trend towards a sex effect on case CNV burden. In a male-only comparison of deletion burden in LoF-intolerant genes, case excess appears stronger (OR=5.25, P=0.08) than in a female-only comparison (OR=1.80, P=0.45). Results from a direct test of burden between male and female cases are consistent with this (2 of 12 males vs. 2 of 89 females, OR=8.36, P=0.07).

## Discussion

We have produced what is, to our knowledge, the first genome-wide study of TTM. While our sample size is limited for a complex psychiatric trait, our results are nonetheless consistent with a genetic contribution to TTM risk and serve as the first meaningful evidence for both common variant and rare variant CNV contributions to this risk. As expected, our TTM GWAS did not identify any genome-wide significant loci. Still, we could leverage a very large and well-powered psychiatric cross-disorder GWAS to demonstrate that cases carry a higher burden of generalized psychiatric polygenic risk. We have made the GWAS summary statistics publicly available (see ***Supplemental Data*** section for download link) in the hope that they will eventually be included in a larger TTM GWAS meta-analysis with sufficient power to identify genome-wide significant risk loci.

We also observed a trend toward an excess of large CNV deletions impacting constrained genes in cases relative to controls. This is consistent with CNV and whole exome sequencing results for conditions known to be comorbid with TTM, such as ADHD, OCD, and MDD.^27–29^ While the overall contribution of rare coding variants to overall TTM genetic risk is not yet established, when studies are sufficiently large, they can be used to pinpoint specific genes whose dysfunction explicitly increases TTM risk.^30^ The rare CNV deletion signal we see in our case/control comparison suggests that we could identify specific TTM risk genes with a sufficiently large sample size.

Taken in full, the results described here suggest that TTM, like other psychiatric conditions, has a genetic architecture consisting of contributions from common and rare variation. Larger sample sizes will be needed to further characterize the genetic architecture of TTM. We hope that the summary statistics shared in this manuscript will be utilized in future studies and that our findings will motivate further analysis of genetic contributions to risk for this understudied condition.

## Data Availability

GWAS summary statistics from a comparison of 101 TTM cases to 488 unaffected controls, in the daner file format: https://figshare.com/s/b94e462100095bf14edb

## Acknowledgments

Dr. Crowley was supported by a grant from the US National Institutes of Health (R01MH110427). Cases were genotyped by the SNP&SEQ Technology Platform in Uppsala, Sweden. The facility is part of the National Genomics Infrastructure (NGI) Sweden and Science for Life Laboratory. The SNP&SEQ Platform is also supported by the Swedish Research Council and the Knut and Alice Wallenberg Foundation. We are grateful for the contributions of our study participants. We thank Stephanie B. Crowley for assistance with the manuscript.

## Supplemental data

GWAS summary statistics from a comparison of 101 TTM cases to 488 unaffected controls, in the ‘daner’ file format: https://figshare.com/s/b94e462100095bf14edb

## References

1. Grant JE, Chamberlain SR. Trichotillomania. Am J Psychiatry. 2016;173(9):868–874.

2. Grant JE, Dougherty DD, Chamberlain SR. Prevalence, gender correlates, and co-morbidity of trichotillomania. Psychiatry Res. 2020;288:112948.

3. Sanderson KV, Hall-Smith P. Tonsure trichotillomania. Br J Dermatol. 1970;82(4):343–350.

4. Galski T. Hair pulling (trichotillomania). Psychoanal Rev. 1983;70(3):331–346.

5. Kerbeshian J, Burd L. Familial trichotillomania. Am J Psychiatry. 1991;148(5):684–685.

6. Schlosser S, Black DW, Blum N, Goldstein RB. The demography, phenomenology, and family history of 22 persons with compulsive hair pulling. Ann Clin Psychiatry. 1994;6(3):147–152.

7. Keuthen NJ, Altenburger EM, Pauls D. A family study of trichotillomania and chronic hair pulling. Am J Med Genet B Neuropsychiatr Genet. 2014;165B(2):167-174.

8. Lenane MC, Swedo SE, Rapoport JL, Leonard H, Sceery W, Guroff JJ. Rates of Obsessive Compulsive Disorder in first degree relatives of patients with trichotillomania: a research note. J Child Psychol Psychiatry. 1992;33(5):925–933.

9. Swedo SE, Rapoport JL. Annotation: trichotillomania. J Child Psychol Psychiatry. 1991;32(3):401–409.

10. Christenson GA, Mackenzie TB, Reeve EA. Familial trichotillomania. Am J Psychiatry. 1992;149(2):283.

11. Novak CE, Keuthen NJ, Stewart SE, Pauls DL. A twin concordance study of trichotillomania. Am J Med Genet B Neuropsychiatr Genet. 2009;150B(7):944–949.

12. Zuchner S, Cuccaro ML, Tran-Viet KN, et al. SLITRK1 mutations in trichotillomania. Mol Psychiatry. 2006;11(10):887–889.

13. Züchner S, Wendland JR, Ashley-Koch AE, et al. Multiple rare SAPAP3 missense variants in trichotillomania and OCD. Mol Psychiatry. 2009;14(1):6–9.

14. Welch JM, Lu J, Rodriguiz RM, et al. Cortico-striatal synaptic defects and OCD-like behaviours in Sapap3-mutant mice. Nature. 2007;448(7156):894–900.

15. Shmelkov SV, Hormigo A, Jing D, et al. Slitrk5 deficiency impairs corticostriatal circuitry and leads to obsessive-compulsive-like behaviors in mice. Nat Med. 2010;16(5):598-602, 1p following 602.

16. Greer JM, Capecchi MR. Hoxb8 is required for normal grooming behavior in mice. Neuron. 2002;33(1):23–34.

17. Shi J, Levinson DF, Duan J, et al. Common variants on chromosome 6p22.1 are associated with schizophrenia. Nature. 2009;460(7256):753–757.

18. Cross-Disorder Group of the Psychiatric Genomics Consortium. Electronic address, Cross-Disorder Group of the Psychiatric Genomics Consortium. Genomic Relationships, Novel Loci, and Pleiotropic Mechanisms across Eight Psychiatric Disorders. Cell. 2019;179(7):1469-1482.e11.

19. Yengo L, Vedantam S, Marouli E, et al. A saturated map of common genetic variants associated with human height. Nature. 2022;610(7933):704–712.

20. International Obsessive Compulsive Disorder Foundation Genetics Collaborative (IOCDF-GC) and OCD Collaborative Genetics Association Studies (OCGAS). Revealing the complex genetic architecture of obsessive-compulsive disorder using meta-analysis. Mol Psychiatry. 2018;23(5):1181–1188.

21. Demontis D, Walters RK, Martin J, et al. Discovery of the first genome-wide significant risk loci for attention deficit/hyperactivity disorder. Nat Genet. 2019;51(1):63–75.

22. Howard DM, Adams MJ, Clarke TK, et al. Genome-wide meta-analysis of depression identifies 102 independent variants and highlights the importance of the prefrontal brain regions. Nat Neurosci. 2019;22(3):343–352.

23. Wray NR, Ripke S, Mattheisen M, et al. Genome-wide association analyses identify 44 risk variants and refine the genetic architecture of major depression. Nat Genet. 2018;50(5):668–681.

24. Lek M, Karczewski KJ, Minikel EV, et al. Analysis of protein-coding genetic variation in 60,706 humans. Nature. 2016;536(7616):285–291.

25. Karczewski KJ, Francioli LC, Tiao G, et al. The mutational constraint spectrum quantified from variation in 141,456 humans. Nature. 2020;581(7809):434–443.

26. Collins RL, Glessner JT, Porcu E, et al. A cross-disorder dosage sensitivity map of the human genome. Cell. 2022;185(16):3041-3055.e25.

27. Satterstrom FK, Walters RK, Singh T, et al. Autism spectrum disorder and attention deficit hyperactivity disorder have a similar burden of rare protein-truncating variants. Nat Neurosci. 2019;22(12):1961–1965.

28. Halvorsen M, Samuels J, Wang Y, et al. Exome sequencing in obsessive-compulsive disorder reveals a burden of rare damaging coding variants. Nat Neurosci. 2021;24(8):1071–1076.

29. Kendall KM, Rees E, Bracher-Smith M, et al. Association of Rare Copy Number Variants With Risk of Depression. JAMA Psychiatry. 2019;76(8):818–825.

30. Weiner DJ, Nadig A, Jagadeesh KA, et al. Polygenic architecture of rare coding variation across 394,783 exomes. Nature. 2023;614(7948):492–499.

