## Supplementary Information for "Genomic Analysis of Trichotillomania"

#### Supplemental Methods

*STAI-Y*

The State-Trait Anxiety Inventory (STAI-Y) is a 40-item self-evaluation questionnaire that measures both state and trait anxiety [1]. The S-Anxiety scale consists of 20 statements that evaluate how respondents feel “right now”, therefore measuring transient anxiety. The T-Anxiety scale consists of 20 statements that assess how people “generally” feel, which measures enduring anxiety. Each item is rated on a 4-point scale ranging from 1-4. The total score is the sum of individual scores (considering that scores should be reversed for certain items) so that the total score for both the S-Anxiety and T-Anxiety scales can vary from 20-80.

*BDI-II*

The Beck Depression Inventory Second Edition (BDI-II) is a 21-item self-report instrument intended to assess the existence and severity of major depression[2]. Each item corresponds to a symptom of depression and is measured on a four-point scale, ranging from 0-3. The total score is the sum of each item score. Total scores ranging from 0-13 indicate no evidence for depression, 14-19 mild to moderate depression, 20-28 moderate to severe depression, and 29-63 severe depression.

*OCI*

The Obsessive-Compulsive Inventory (OCI) is a 42-item self-report instrument for determining the diagnosis and severity of OCD [3], which was designed for both clinical and non-clinical populations. The OCI encompasses more of the heterogeneity of OCD by measuring seven subscales: Washing, Checking, Doubting, Ordering, Obsessing, Hoarding, and Mental Neutralizing. Each item is rated on a 0-4 Likert scale and captures both the symptom frequency and associated distress.

*Phenotypic analysis*

Pedigree structure and family history reports were examined for the majority of probands with TTM (N=90) for whom the relevant data was available. We recorded the frequency of comorbid psychiatric conditions in the probands and their first-degree relatives. We also report frequencies of the three most common comorbidities by gender. Statistical analysis was performed using contingency tables and logistic regression in SAS version 9.1 (SAS Institute, Cary, NC).

Scores from the STAI, BDI, and OCI were analyzed for probands when available. Means and frequencies were obtained using PROC MEANS and PROC FREQ, respectively, in SAS version 9.1.

*Recruitment and selection of controls for genetic study*

Unaffected control subjects included in this study were initially recruited by Knowledge Networks via random-digit dialing of residential phone numbers. Participants were pointed to a website to learn more about the study, given preliminary online informed consent, and completed a self-report clinical assessment. The online assessment included the Composite International Diagnostic Interview-Short Form (CIDI-SF), modified for lifetime common mood, anxiety, and psychosis or bipolar disorder. We excluded individuals who failed to complete the CIDI-SF or endorsed schizophrenia, schizoaffective disorder, auditory hallucinations, delusions, or bipolar disorder.

*Merging case and control genotype data*

For downstream analyses, we merged case and control genotype data on markers in both datasets. We identified 633,201 markers with matching IDs and human reference build GRCh37 genomic coordinates (chromosome and position). Of these, we identified a total of 575,596 markers that had genotype missingness < 0.02 in both the case-only and control-only sample groups. Finally, we required that SNPs have a case/control missingness Fisher's exact test two-sided p-value >= 0.01 and that for concordance checks, samples have a reported phenotype (case/control) and sex (male/female). After placing these constraints on the data, our merged set contained 575,579 markers across 116 cases and 1033 controls.

*Pre-analysis quality control (QC)*

For the pre-analysis QC of genotype array data, we utilized the preimp_dir function from the Ricoipili pipeline (version: 2019_Jun_25.001) [4], which was designed to streamline the setup and execution of GWAS. Default parameters were used, which include: marker call rate >= 0.95; sample call rate >= 0.98; sample FHET +/- 0.2; reject observed/expected sex mismatch; reject invariant markers; control HWE P >= $1x10^{-6}$; case HWE P >= $1x10^{-10}$. We were left with 491,616 markers across 116 cases and 985 controls.

*Biological QC*

We selected samples for inclusion in all downstream analyses based on two criteria: 1) samples needed to be unrelated, and 2) samples had to be of European ancestry since this is the predominant self-declared ancestry of cases, and it is important to avoid population stratification in setting up analyses. We obtained 1000 Genomes Phase 3 genotype data and merged these data with our case/control data across a total of 109,964 shared SNPs. We conducted PCA using PLINK v1.90b3.45 and computed the mean and standard deviation across PCs 1-4 for 1000 genomes EUR samples. We kept a case/control cohort sample if it fell within 10 standard deviations of the EUR mean across PCs 1-4 (107 cases and 495 controls total). We also conducted relatedness pruning, controlling for instances of sample duplication or cryptic relatedness, and removed four control samples. As a final control against ancestry outliers in the data biasing results, we trimmed any samples from our analysis that fell outside of 6 standard deviations from the mean for PC1 or PC2. This left us with 589 homogenous European samples (101 cases, 488 controls) usable for subsequent case/control comparisons.

*Imputation and GWAS*

For imputation and genome-wide association study (GWAS) of common variation, we again utilized the Ricoipili pipeline (version: 2019_Jun_25.001) [4], specifically the functions ‘impute_dirsub’ and ‘postimp_navi’, respectively. Imputation was carried out using 1000 genomes phase 3 genotypes, with eagle v2.3.5 [5] being used for pre-phasing and minimac3 v2.0.1 [6] being used for imputation. The function ‘pcaer_sub’ was used to conduct PCA on LD-pruned genotype data and produce the first 20 PCs across samples. Only PC1 was found to be associated with sample case status, and this, along with sex, was used as a covariate for the GWAS.

*Polygenic risk score calculation*

All polygenic risk scores (PRS) were calculated using PRS-CS (version: Sept 10, 2020) [7], a Bayesian method that enables PRS prediction without p-value thresholding by using available ancestry-matched patterns of linkage disequilibrium. We utilized the 1000 genomes phase 3 European reference panel compiled by PRS-CS authors (https://github.com/getian107/PRScs). For all PRS calculations, we focused on a set of post-imputation SNPs that met the following criteria in PLINK: SNP missingness < 0.1, individual missingness < 0.1, MAF > 0.05, Hardy-Weinberg p-value > $1x10^{-6}$, and excluded the major histo-compatibility region (MHC). For PRS calculation, we only used SNPs in training data if they were unambiguous (i.e., non-A/T and non-G/C SNPs).

*Polygenic risk score analysis*

All association tests between PRS and TTM case status were conducted using R v3.4.3. Before analysis, each PRS was standardized, and all values described and plotted here refer to the PRS ‘standard score’. For each association test, a logistic regression model was formed with case status as the outcome and standardized PRS as the critical predictor. As with GWAS, PC1 and sex were utilized as covariates in these models. Any significant results were repeated in a subset of the cohort pruned of samples with PC1<0.05 (case/control N=80/457) to ensure that samples with PC1>=0.05 are not driving the case/control difference in PRS. No results described in our manuscript showed discordant results in the PC1-pruned comparison.

*CNV calling*

We called copy number variants from TTM cases and unrelated controls using PennCNV v1.0.4 [8]. Calling was done using the PennCNV script ‘detect_cnv.pl’, providing as input the provided HMM file ‘hhall.hmm’, a GC model file for SNP markers produced via the command ‘cal_gc_snp.pl’, a pfb file produced across case/control intensity data, and intensity metrics (B allele frequency, Log R ratio) per SNP marker/sample combination. Contiguous segments were merged via iterative calling of the ‘clean_cnv.pl’ function ‘combineseg’ using default settings, until no additional mergers of contiguous calls in individual samples occurred. Only CNV calls at least 100kb in length and overlapping at least 15 SNP markers were retained for analysis.

*CNV QC*

CNV analyses focused on the same samples used in the GWAS, with additional subsetting to ensure a well-controlled case/control comparison. Specifically, we retained case and control samples that met the following criteria: *n* raw CNV calls <= 20; *n* basepairs overlapping raw CNV calls <= 20,000,000; LRR_SD < 0.24; BAF_DRIFT < 0.001; and absolute value of waviness factor < 0.04. Thresholding for the burden of raw CNVs was selected manually, while burden on intensity metrics was set based on ideal thresholds described in Huang et al. 2017 [9]. We were left with 101 cases and 462 controls for the CNV analysis.

*CNV filtering*

We adapted the code and protocol for CNV filtration from a recent meta-analysis involving CNVs of similar minimum sizes [10]. We defined blacklist loci using the following set of annotations: 1) predefined telomere and centromere regions, with 500kb of padding; 2) poly-N regions in the reference genome GRCh37, derived using (https://github.com/lh3/seqtk); 3) segmental duplication loci [11] 4) simple repeats, low complexity regions and satellite regions from RepeatMasker (RepeatMasker Open-3.0); 5) immunoglobulin and T-cell receptor genes; and 6) CNV loci associated with LCL status in EBV-transformed cell-lines [12]. We removed any CNV call where over 50% of the call locus overlapped the union of the loci described above. We also removed CNVs that were found at a frequency of 0.01 or greater in one of the following: 1) case data, 2) control data, 3) the combined case/control cohort, and 4) gnomAD v2.1 "non-neuro" cohort structural variant callset data and all defined population subsets [13]. After CNV filtration, across 563 samples, we observe 153 deletions and 162 duplications. These correspond to CNV rates of 0.27 deletions and 0.29 duplications per sample.

*CNV analysis*

All statistical analyses of CNV burden were done in R v4.2.2. We tested the basic hypothesis that TTM cases have an excess of CNV deletions and duplications, specifically within genes constrained across human populations. We defined 'constrained' genes as those with a probability of loss of function intolerance (pLI) > 0.9 (n=3,063 genes total) [14,15] (PMIDs 27535533, 35396579). Genes with pLI <= 0.9 (n=16,095) were considered not constrained in the general population. We tested the burden of deletions and duplications separately, and for each one, 1) built 2x2 contingency tables for males and females for carriers and non-carriers in case and control groups, and 2) meta-analyzed the sex-stratified tables using a one-sided Cochran-Mantel-Haenszel Exact Test.

#### References

1. Spielberger, C.D. (2012). State-Trait Anxiety Inventory for Adults. (American Psychological Association). 10.1037/t06496-000 10.1037/t06496-000.
2. Beck, A.T., Steer, R.A., and Brown, G. (2011). Beck Depression Inventory–II. (American Psychological Association). 10.1037/t00742-000 10.1037/t00742-000.
3. Foa, E.B., Kozak, M.J., Salkovskis, P.M., Coles, M.E., and Amir, N. (1998). The validation of a new obsessive–compulsive disorder scale: The Obsessive–Compulsive Inventory. Psychological Assessment *10*, 206–214. 10.1037/1040-3590.10.3.206.
4. Lam, M., Awasthi, S., Watson, H.J., Goldstein, J., Panagiotaropoulou, G., Trubetskoy, V., Karlsson, R., Frei, O., Fan, C.-C., De Witte, W., et al. (2019). RICOPILI: Rapid Imputation for COnsortias PIpeLIne. Bioinformatics. 10.1093/bioinformatics/btz633.
5. Loh, P.-R., Danecek, P., Palamara, P.F., Fuchsberger, C., A Reshef, Y., K Finucane, H., Schoenherr, S., Forer, L., McCarthy, S., Abecasis, G.R., et al. (2016). Reference-based phasing using the Haplotype Reference Consortium panel. Nat Genet *48*, 1443–1448. 10.1038/ng.3679.
6. Das, S., Forer, L., Schönherr, S., Sidore, C., Locke, A.E., Kwong, A., Vrieze, S.I., Chew, E.Y., Levy, S., McGue, M., et al. (2016). Next-generation genotype imputation service and methods. Nat Genet *48*, 1284–1287. 10.1038/ng.3656.
7. Ge, T., Chen, C.-Y., Ni, Y., Feng, Y.-C.A., and Smoller, J.W. (2019). Polygenic prediction via Bayesian regression and continuous shrinkage priors. Nat Commun *10*, 1776. 10.1038/s41467-019-09718-5.
8. Wang, K., Li, M., Hadley, D., Liu, R., Glessner, J., Grant, S.F.A., Hakonarson, H., and Bucan, M. (2007). PennCNV: an integrated hidden Markov model designed for high-resolution copy number variation detection in whole-genome SNP genotyping data. Genome Res. *17*, 1665–1674. 10.1101/gr.6861907.
9. Huang, A.Y., Yu, D., Davis, L.K., Sul, J.H., Tsetsos, F., Ramensky, V., Zelaya, I., Ramos, E.M., Osiecki, L., Chen, J.A., et al. (2017). Rare Copy Number Variants in NRXN1 and CNTN6 Increase Risk for Tourette Syndrome. Neuron *94*, 1101-1111.e7. 10.1016/j.neuron.2017.06.010.
10. Collins, R.L., Glessner, J.T., Porcu, E., Lepamets, M., Brandon, R., Lauricella, C., Han, L., Morley, T., Niestroj, L.-M., Ulirsch, J., et al. (2022). A cross-disorder dosage sensitivity map of the human genome. Cell *185*, 3041-3055.e25. 10.1016/j.cell.2022.06.036.
11. Bailey, J.A., Gu, Z., Clark, R.A., Reinert, K., Samonte, R.V., Schwartz, S., Adams, M.D., Myers, E.W., Li, P.W., and Eichler, E.E. (2002). Recent segmental duplications in the human genome. Science *297*, 1003–1007. 10.1126/science.1072047.
12. Shirley, M.D., Baugher, J.D., Stevens, E.L., Tang, Z., Gerry, N., Beiswanger, C.M., Berlin, D.S., and Pevsner, J. (2012). Chromosomal variation in lymphoblastoid cell lines. Hum Mutat *33*, 1075–1086. 10.1002/humu.22062.
13. Collins, R.L., Brand, H., Karczewski, K.J., Zhao, X., Alföldi, J., Francioli, L.C., Khera, A.V., Lowther, C., Gauthier, L.D., Wang, H., et al. (2020). A structural variation reference for medical and population genetics. Nature *581*, 444–451. 10.1038/s41586-020-2287-8.
14. Lek, M., Karczewski, K.J., Minikel, E.V., Samocha, K.E., Banks, E., Fennell, T., O'Donnell-Luria, A.H., Ware, J.S., Hill, A.J., Cummings, B.B., et al. (2016). Analysis of protein-coding genetic variation in 60,706 humans. Nature *536*, 285–291.

10.1038/nature19057.

15. Singh, T., Poterba, T., Curtis, D., Akil, H., Al Eissa, M., Barchas, J.D., Bass, N., Bigdeli, T.B., Breen, G., Bromet, E.J., et al. (2022). Rare coding variants in ten genes confer substantial risk for schizophrenia. Nature *604*, 509–516. 10.1038/s41586-022-04556-w.

#### Figure S1: PCA plot for a subset of individuals with European ancestry

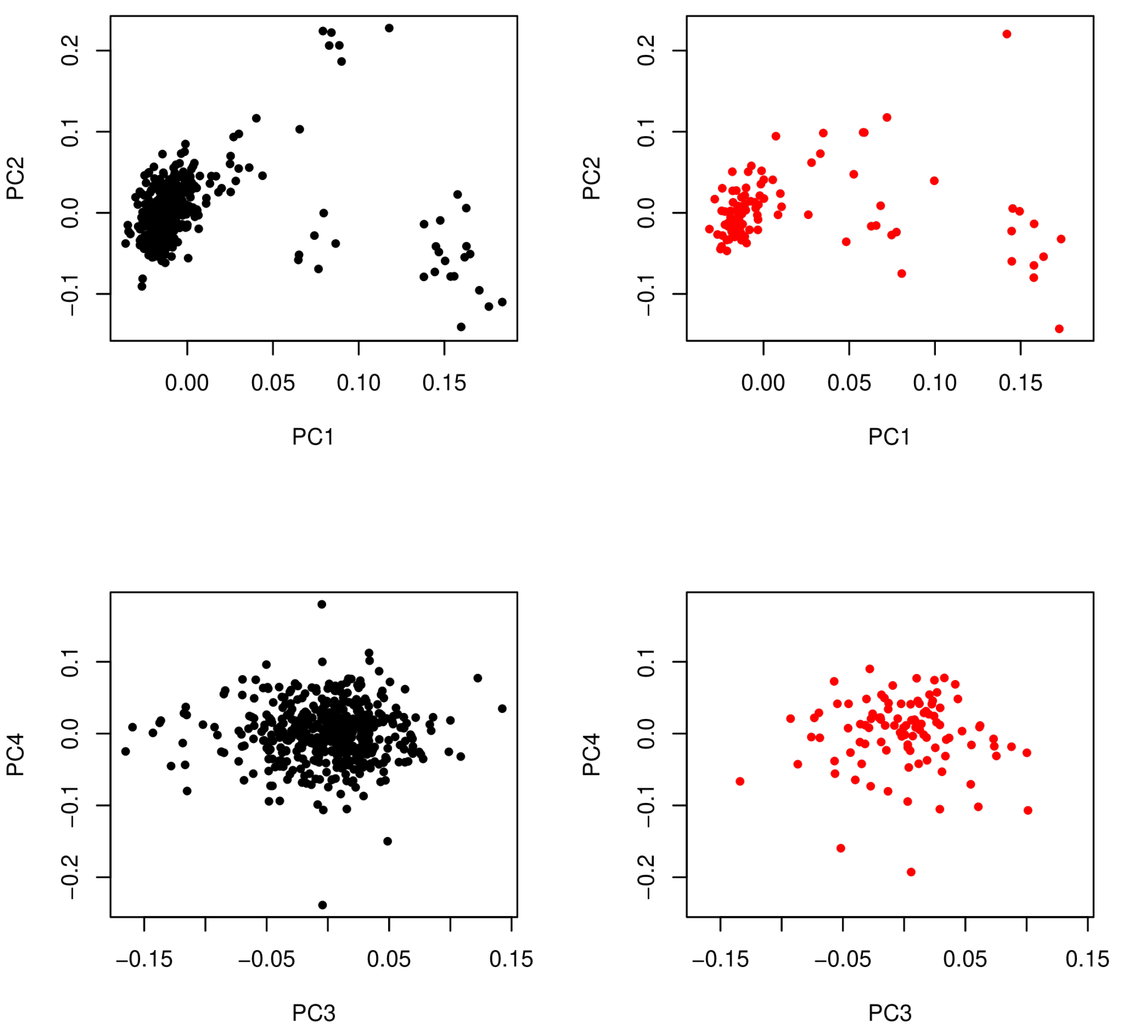

Figure S1 shows the results of the principal component analysis of 109,964 LD-pruned common variants across 101 European cases and 488 ancestry-matched controls. We show the first 4 PCs here because they explain the large majority of variance across the first 20 PCs. The first PC is the only one with a detectable difference (logistic regression $P < 0.01$) between cases and controls and was used as a covariate in subsequent GWAS and PRS analyses.

#### Figure S2: QQ plot for GWAS of European TTM cases and ancestry-matched controls

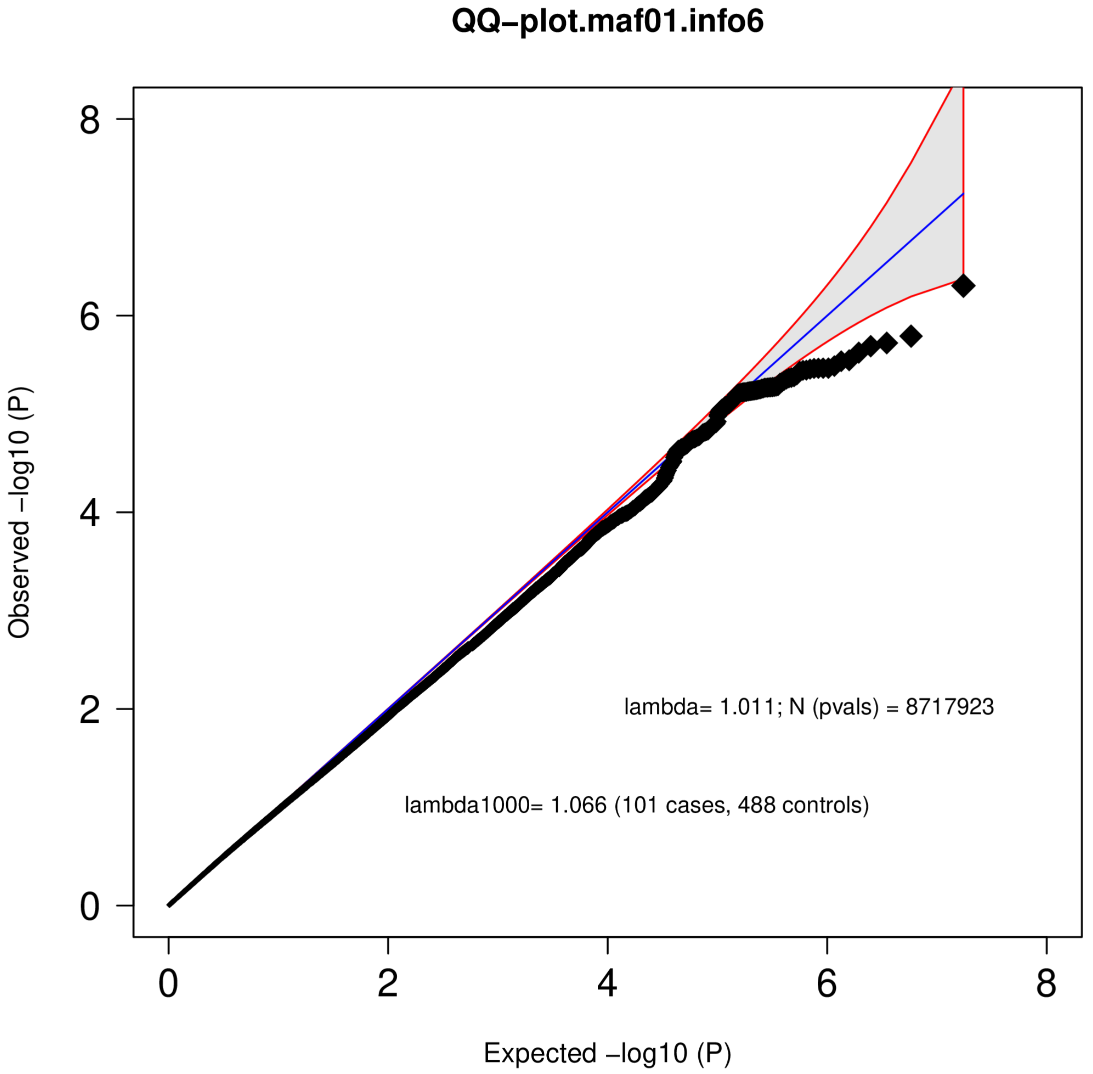

Figure S2 shows a quantile-quantile plot of observed log-transformed P-values versus expected log-transformed P-values across the GWAS of 101 European-ancestry TTM cases and 488 population-matched controls. A total of 8,717,923 tests were performed here, and across these tests, we find no evidence for P-value inflation or deflation based on a lambda of 1.011.

#### Figure S3: Manhattan plot from GWAS of TTM cases and unaffected controls

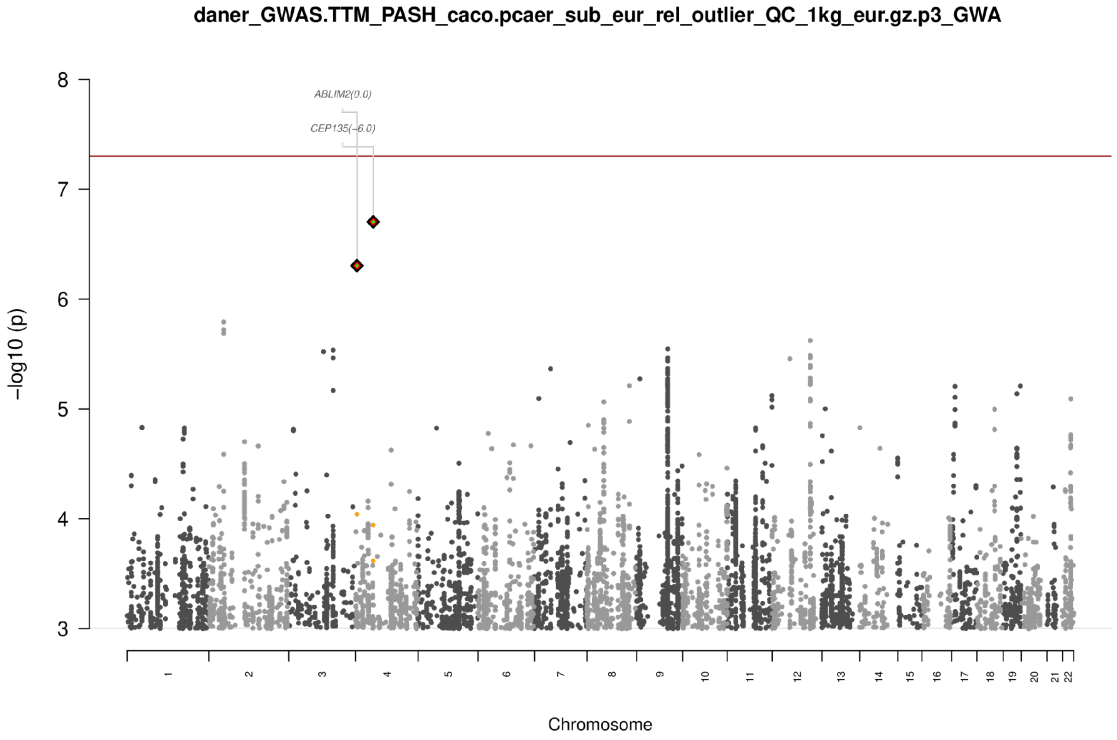

Figure S3 depicts a Manhattan plot of TTM GWAS summary statistics across 8,717,923 common variants. We do not observe any individual SNPs that pass the genome-wide significance threshold ($5x10^{-8}$, or -log10p=7.3).

#### Figure S4: PRS case/control comparisons (female-only, male-only)

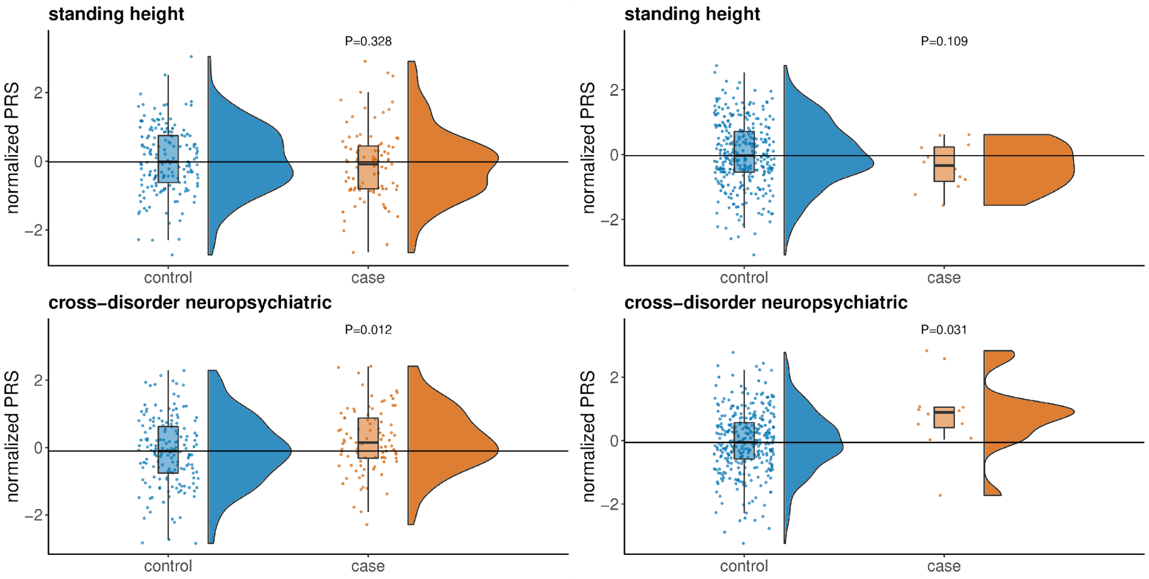

Figure S4 depicts the distribution of normalized PRS for standing height and cross-disorder neuropsychiatric conditions, stratified into a female-only comparison (left, case/control n = 89/165) and a male-only comparison (right, n=12/323). Both female-only and male-only comparisons have detectable (T-test two-sided $P<0.05$) differences in normalized PRS for neuropsychiatric conditions between cases and controls, but the increase in psychiatric PRS for male cases versus male controls appears higher than in female cases versus female controls.

#### Figure S5: Deletion CNVs (*NRXN1* gene, *RBFOX1* gene)

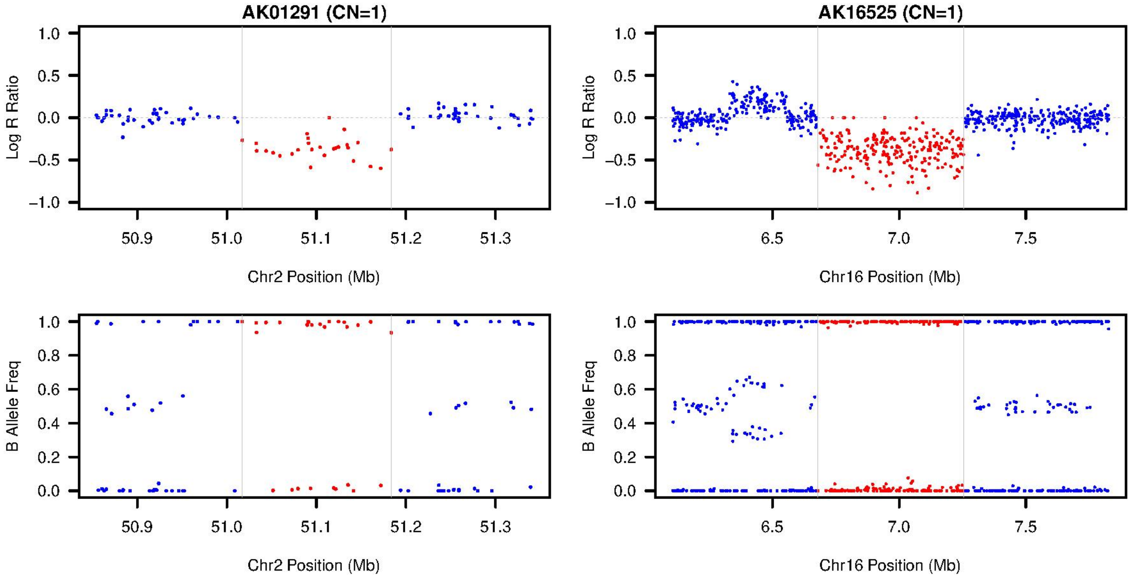

Figure S5 shows the distribution of B allele frequency (BAF) and Log-R Ratio (LRR) values across call sites for a deletion overlapping *NRXN1* (left, hg19 coordinates = chr2:51017206-51183559) and a deletion overlapping *RBFOX1* (right, hg19 coordinates = chr16:6680132-7253942), both of which were detected in separate TTM cases. Both pass visual inspection because the BAF and LRR distributions are consistent with a copy number of 1 within the call sites. It is worth noting that next to the *RBFOX1* deletion, there appears to be a neighboring duplication upstream, leading to a copy state of 3.

### Figure S6: Deletion CNVs (15q11.2 locus, *CSMD1* gene)

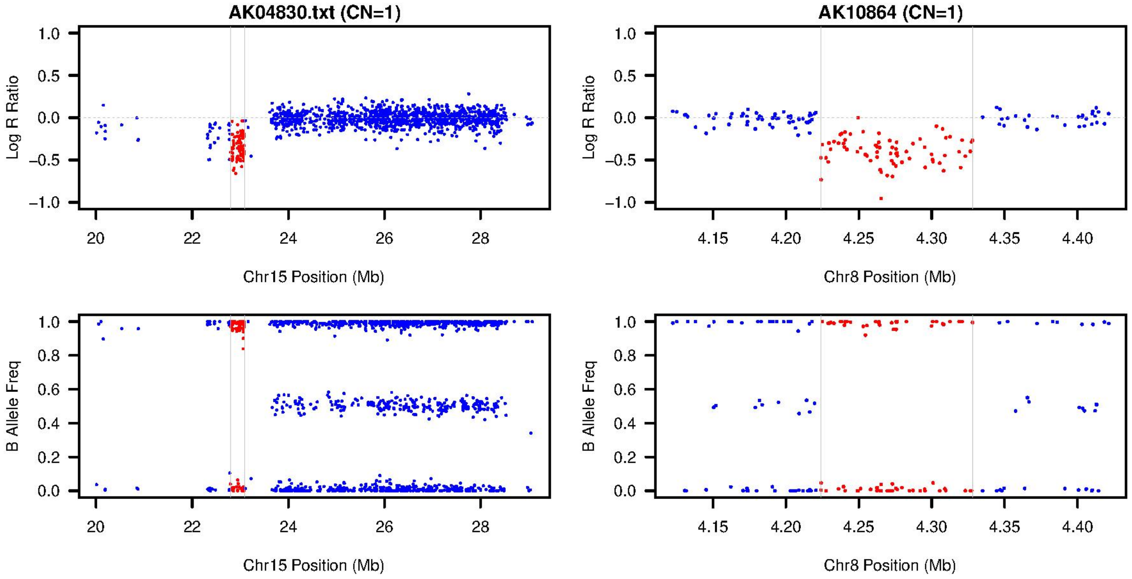

Figure S6 shows BAF and LRR values across call sites for a 15q11.2 deletion (left, hg19 coordinates = chr15:22794494-23086929) and a deletion overlapping *CSMD1* (right, hg19 coordinates = chr8:4224080-4327992), both of which were detected in separate TTM cases. As with the BAF/LRR plots shown in Figure S5, both pass visual inspection and are consistent with a copy number of 1 within each call site.

#### Table S1. Frequency of comorbidities in probands and first-degree relatives

| Comorbidity | Percentage (Probands) | Percentage (first-degree relatives) |
|---|---|---|
| TTM | 100% | 10.7% |
| Mood disorders | 33.3% | 14.1% |
| Anxiety disorders | 30.0% | 14.6% |
| OCD | 12.2% | 2.9% |
| Substance Abuse | 8.9% | 12.5% |
| AD/HD | 7.8% | 1.8% |
| Eating disorders | 6.7% | 1.8% |
| Migraines | 3.3% | 2.4% |
| Tourette's syndrome/tics | 3.3% | 0.5% |
| Obesity | 2.2% | 2.9% |
| Seizures | 2.2% | 1.6% |
| Body Dysmorphic Disorder | 1.1% | 0.3% |
| Cutters | 1.1% | 0.0% |
| Kleptomania | 1.1% | 1.0% |
| Learning disabilities | 1.1% | 2.1% |
| Sleep disorders | 1.1% | 0.3% |
| Suicide/suicide attempts | 1.1% | 0.3% |

#### Table S2. Frequencies of related disorders by sex

| Comorbidity | Percentage (Females) | Percentage (Males) | Fisher's exact p-value |
|---|---|---|---|
| Mood disorders | 32.9% | 37.5% | 1 |
| Anxiety disorders | 31.7% | 12.5% | 0.43 |
| OCD | 13.4% | 0% | 0.59 |

#### Table S3. BDI-II results for TTM cases (N = 51)

| Category | Frequency | Percentage |
|---|---|---|
| not depressed (0-13) | 28 | 54.9% |
| mild-moderate depression (14-19) | 10 | 19.6% |
| moderate-severe depression (20-28) | 6 | 11.8% |
| severe depression (29-63) | 7 | 13.7% |

#### Table S4. OCI results

| Subscale (# items) | Type | Mean | SD | Min | Max | N |
|---|---|---|---|---|---|---|
| Checking (9) | Frequency | 0.62 | 0.61 | 0 | 2.56 | 44 |
| | Distress | 0.39 | 0.51 | 0 | 2.33 | 43 |
| Hoarding (3) | Frequency | 0.94 | 0.89 | 0 | 3.00 | 44 |
| | Distress | 0.62 | 0.85 | 0 | 3.33 | 43 |
| Neutralizing (6) | Frequency | 0.66 | 0.72 | 0 | 3.67 | 44 |
| | Distress | 0.50 | 0.67 | 0 | 3.83 | 42 |
| Obsessing (8) | Frequency | 1.06 | 0.76 | 0 | 3.00 | 43 |
| | Distress | 1.16 | 0.93 | 0 | 3.13 | 41 |
| Ordering (5) | Frequency | 1.26 | 1.00 | 0 | 4.00 | 44 |
| | Distress | 0.86 | 0.92 | 0 | 4.00 | 42 |
| Washing (8) | Frequency | 0.79 | 0.87 | 0 | 3.63 | 42 |
| | Distress | 0.57 | 0.83 | 0 | 3.75 | 41 |
| Doubting (3) | Frequency | 0.92 | 1.07 | 0 | 3.67 | 44 |
| | Distress | 0.74 | 0.95 | 0 | 3.33 | 41 |
| Total (42) | Frequency | 0.85 | 0.66 | 0 | 3.14 | 41 |
| | Distress | 0.68 | 0.65 | 0 | 3.24 | 37 |

#### Table S5. STAI results

| Subscale | Mean | Std Dev | Min | Max | N |
|---|---|---|---|---|---|
| S-anxiety | 43.26 | 11.85 | 21 | 69 | 53 |
| T-anxiety | 48.06 | 11.97 | 23 | 76 | 54 |
